# Reduced risk of placental parasitaemia associated with complement fixation on *Plasmodium falciparum* by antibodies among pregnant women

**DOI:** 10.1101/2021.06.02.21258254

**Authors:** D. Herbert Opi, Michelle J. Boyle, Alistair McLean, Linda Reiling, Jo-Anne Chan, Danielle I. Stanisic, Alice Ura, Ivo Mueller, Freya J. Fowkes, Stephen J. Rogerson, James G. Beeson

## Abstract

**Background:** The pathogenesis of malaria in pregnancy (MiP) involves accumulation of *P. falciparum*-infected red blood cells (pRBCs) in the placenta, contributing to poor pregnancy outcomes. Parasite accumulation is primarily mediated by *P. falciparum* erythrocyte membrane 1 (PfEMP1). Magnitude of IgG to pRBCs has been associated with reduced risk of MiP in some studies, but associations have been inconsistent. Further, antibody effector mechanisms are poorly understood, and the role of antibody complement interactions is unknown.

**Methods:** Studying a longitudinal cohort of pregnant women (n=302) from a malaria-endemic province in Papua New Guinea (PNG), we measured the ability of antibodies to fix and activate complement using placental binding pRBCs and PfEMP1 recombinant domains. We determined antibody-mediated complement inhibition of pRBC binding to the placental receptor, chondroitin sulfate A (CSA) and associations with protection against placental parasitaemia.

**Results:** Some women acquired antibodies that effectively promoted complement fixation on placental-binding pRBCs. Complement fixation correlated with IgG1 and IgG3 antibodies, which dominated the response. There was, however, limited evidence for membrane attack complex activity or pRBC lysis or killing. Importantly, a higher magnitude of complement fixing antibodies was prospectively associated with reduced odds of placental infection at delivery. Using genetically-modified *P. falciparum* and recombinant PfEMP1 domains, we found that complement-fixing antibodies primarily targeted a specific variant of PfEMP1 (known as VAR2CSA). Furthermore, complement enhanced the ability of antibodies to inhibit pRBC binding to CSA, which was primarily mediated by complement C1q protein.

**Conclusion:** These findings provide new insights into mechanisms mediating immunity to MiP and reveal potential new strategies for developing malaria vaccines that harness antibody-complement interactions.

## INTRODUCTION

Malaria in pregnancy (MiP) causes significant maternal, fetal and neonatal mortality and is a major health issue globally (1). Parasite accumulation in the placenta is a key feature of MiP following infection with *Plasmodium falciparum* (2, 3) but is not prominent with other human infecting *Plasmodium* species. This largely results from the selective binding of pRBCs to chondroitin sulfate A (CSA) expressed on syncytiotrophoblasts (4, 5), and other binding interactions may play secondary roles (6, 7). This is mediated by a *Plasmodium falciparum* erythrocyte membrane protein 1 (PfEMP1) variant surface antigen, VAR2CSA, encoded by the *var* multigene family (5, 8). The risk of MiP is greatest in primigravid women, and generally decreases with successive pregnancies in malaria endemic areas due in part to the acquisition of protective antibodies directed against placental-binding *P. falciparum* infected red blood cells (pRBCs)(9). Antibodies to placental-binding pRBCs and VAR2CSA have been associated with improved outcomes in some studies, although associations have not been entirely consistent (10). How antibodies to pRBCs and VAR2CSA function in protective immunity to MiP and improved birth outcomes is not fully understood (10), and these key knowledge gaps, are restricting advancement of MiP vaccines. Existing data shows that antibodies may act via inhibition of placental adhesion of pRBCs (11) and promoting phagocytosis of pRBCs (12). However, a recent systematic review found that data on associations between these antibody functions and protection from the consequences of MiP are limited and variable (10) and data suggest these mechanisms may not fully explain immunity to MiP. VAR2CSA is a leading vaccine candidate for MiP with two VAR2CSA based vaccines having completed phase I trials (13, 14), which highlights the importance of a strong understanding of immunity to inform further vaccine design and development for MiP.

Antibodies to placental-binding pRBCs and VAR2CSA are dominated by IgG1 and IgG3 subclasses (15, 16), which have the potential to fix and activate human complement against infecting pathogens (17). Complement activation via the classical pathway is initiated by binding of complement C1q to antigen-antibody complexes. This leads to a cascade of activation of other complement components including C3 and culminates in the formation of the C5-9 membrane attack complex (MAC)(18). Complement fixation can mediate protective functions through several mechanisms: i) MAC formation can lead to cell lysis or death, ii) complement components C3 and C5 can promote phagocytosis by monocytes and neutrophils through interactions with complement receptors expressed on those cells, iii) binding of complement components to a pathogen surface may also have direct inhibitory or neutralizing activity (18-20). Antibody-mediated complement fixation against *P. falciparum* has been implicated in immunity in non-pregnant individuals, targeting merozoites (21, 22), sporozoites (23, 24) and gametocytes (25). While there is a potential role for antibody-mediated complement fixation in immunity against MiP, this has not been established and RBCs are known to express complement regulatory proteins that can inhibit complement activation and confer resistance to lysis (26). Older studies have reported antibody-mediated complement fixation on the surface of mature pigmented trophozoite stage pRBCs, but these did not assess different functional activities or associations with protection (26, 27). Further, MiP has also been associated with excessive complement activation and production of inflammatory products mediating adverse outcomes (28). Therefore, the role of complement fixation in preventing or reducing placental infection remains unclear, and there has been little investigation of its potential role in immunity.

We hypothesised that acquired antibodies against VAR2CSA in some pregnant women may fix complement on placental binding pRBCs, which may contribute to the control or prevention of placental parasitaemia. Using a prospective longitudinal cohort study of malaria-exposed pregnant women from PNG, we investigated antibody-complement interactions in immunity to MiP. We investigated antigenic targets of complement fixation on the surface of pRBCs using genetically-modified pRBCs and recombinant VAR2CSA proteins, and evaluated functional mechanisms of complement-fixing antibodies. We examined whether antibody-mediated complement fixation on pRBCs is associated with reduced risk of placental infection.

## MATERIALS AND METHODS

### Study population

This study was part of a larger prospective longitudinal cohort study looking at the risk factors for malaria and adverse birth outcomes among pregnant women, described elsewhere (29). The study was carried out in the malaria-endemic province of Madang in Papua New Guinea (PNG), September 2005 to October 2007. 470 pregnant women >16 years of age attending their first antenatal care visit at the Alexishafen Health Centre were recruited into the study following informed voluntary consent. Women were followed up at 30-34 weeks gestation and at delivery. At enrolment, women received chloroquine (9 or 12 tablets, 150 mg base) and (when available) sulphadoxine pyrimethamine (500/25 mg, three tablets), followed by weekly chloroquine prophylaxis (two 150 mg tablets weekly), and ferrous sulphate 270 mg and folic acid 0.3 mg daily, according to local guidelines. Adherence to prophylaxis was, however, not monitored. Inclusion criteria included no history of multiple births (for example past delivery of twins) and delivery complications, intention to deliver at the Alexishafen Health Centre, haemoglobin (Hb) >5g/dl and evidence of fetal movement. This study is restricted to 302 women with paired data available for enrolment and delivery visits. During each visit, clinical and demographic data were recorded. Peripheral blood samples were collected at each visit and plasma and serum samples separated and frozen. Placental blood and placental biopsy samples were collected at delivery if the delivery occurred at the clinic (n=233). At each visit, peripheral parasitaemia was determined by microscopy on thick and thin blood films and *Plasmodium spp* infection confirmed by PCR (29). Placental infection was determined by histology on fixed Giemsa-stained placental sections by light microscopy and was classified as no-infection, past infection, chronic infection or active infection as previously described (30, 31). The human genetic polymorphisms of South-East Asian Ovalocytosis (SAO), Complement Receptor 1 (CR1) and α+thalassaemia are common in this population and have been associated with protection against severe clinical malaria in some studies ((32, 33)). Therefore, these were included as possible confounders in our study. Molecular typing was conducted as previously described (34). Samples from malaria non-exposed residents of Melbourne, Australia were used as negative controls in all assays.

### Parasite culture

For all assays, CS2 *P. falciparum* parasite strain was used. CS2 binds to CSA and predominantly expresses the *var* gene *var2csa* (4, 35). Assays on the role of PfEMP1 included a transgenic CS2 *P. falciparum* isolate in which the genetic deletion of the PfEMP1 trafficking protein skeleton-binding protein 1, SBP1 knock-out (CS2-SBP1KO), significantly impairs PfEMP1 RBC surface expression (36). Additional experiments were carried out using the XIE *P. falciparum* isolate that originated from a pregnant PNG woman (37). XIE *P. falciparum* was adapted to *in vitro* culture and selected for adhesion to immobilized CSA giving rise to the XIE-CSA isolate that expresses *var2csa* as the dominant transcript (37). Parasites were cultured in human blood group O RBCs and RPMI-HEPES medium (Thermo Fisher Scientific) supplemented with 5% heat-inactivated human serum (vol/vol) from a pool of malaria non-exposed Australian Red Cross donors and 0.25% AlbuMAX™ II (Thermo Fisher Scientific) (vol/vol), NaHCO_3_ and gentamicin (complete culture medium). The parasites were then maintained in a gas mixture of 1% O_2_, 4% CO_2_ and 95% N_2_ at 37°C. Knobby pRBCs were selected for and maintained by regular flotation in 0.75% gelatin in RPMI-HEPES.

### Adhesion inhibition assays

The ability of immune antibodies in the presence or absence of complement to inhibit pRBC adhesion to CSA was assessed using a modified version of a static-based binding assay described previously (38). Petri dishes were coated with 2.5μg/ml of CSA diluted in PBS and incubated overnight at 4°C. Plates were subsequently blocked with 1% bovine serum albumin (BSA) in PBS followed by gentle washing. Mature pigmented-trophozoite stage CS2 pRBCs at approximately 3% parasitaemia and 1% haematocrit, in RPMI, were opsonized with a pool of antibodies of serum samples from either PNG pregnant women (N=9) with high IgG reactivity to VAR2CSA-DBL5 (3D7) as determined by ELISA or malaria non-exposed Australian donors (N=10). The pooled samples were tested in assays at a final concentration of 10% in RPMI-HEPES. Additionally, to test for complement fixation, samples were concurrently incubated with normal serum (NS; complement active) or heat inactivated serum (HIS; complement inactive) at a final dilution of 25% in RPMI, 10μg/ml purified human C1q (Millipore) or C1q-depleted human serum (Millipore) at a final dilution of 10% in RPMI, or RPMI (negative control) for 30 minutes at 37°C at a final volume of 50μl. Both NS and HIS were from malaria non-exposed Australian donors. The parasite suspension was then added onto the CSA coated spots and incubated at 37°C for a further 15 minutes. Unbound cells were washed off with gentle agitation and bound pRBCs were fixed in 2% glutaraldehyde in PBS followed by staining with 10% Giemsa. Each sample was tested in duplicate spots and repeated in 3-6 independent assays. Images of adherent pRBCs were captured using an inverted microscope with 8 images captured for each of the duplicate spots for each protein. Adherent pRBCs were counted and the results expressed as the mean number of pRBCs bound per mm^2^.

### Recombinant proteins

For ELISA-based assays, antibody levels were determined for 3 VAR2CSA recombinant proteins representing 2 allelic variants; DBL5 (3D7), DBL3 (7G8) and DBL5 (7G8). All recombinant proteins were cloned and produced in *Pichia pastoris* (39). DBL5 and DBL3 recombinant proteins were selected because compared to other VAR2CSA domains they are highly immunogenic in natural infections and elicit some degree of cross-reactive and adhesion-blocking antibodies (8, 40-44), and also promote opsonic phagocytosis by monocytes (45).

### Plate-based complement fixation assays with recombinant proteins

96-well plates were coated with 1μg/ml of VAR2CSA-DBL5 (3D7) or VAR2CSA-DBL3 (7G8) recombinant proteins and incubated at 4°C overnight. Plates were then blocked with 2% casein in PBS at 37°C for 2 hours and then incubated with antibodies at a dilution of 1/100 followed by purified C1q (Millipore) at 10μg/ml or C5-depleted serum (Millipore) at a dilution of 1/10 for the detection of C3, at room temperature for 1 hour. A combination of goat anti-C1q plus HRP-conjugated rabbit anti-goat antibodies and rabbit anti-C3 plus HRP-conjugated goat anti-rabbit at dilutions of 1/2000 were used for the detection of C1q and C3 respectively. Reactivity was determined by measuring the optical density (OD) at 405nm following the addition of ABTS (2,2’-azino-bis(3-ethylbenzothiazoline-6-sulfonic acid)) and stopping of the reaction with 1% sodium dodecyl sulfate (SDS) after 15 minutes to 1 hour. Blank wells, without antibodies added, were used to subtract non-specific signal from each well. IgG to VAR2CSA was measured using goat anti-human IgG-HRP (Millipore) while IgG subclasses to VAR2CSA were measured using sheep anti-human IgG1, IgG2, IgG3, IgG4 (HRP) antibodies (Binding site). Optical density (OD) results for each assay plate were then standardized to account for plate-to-plate variation by using values from five PNG positive control serum samples that were included on every plate. The positive control samples were from individuals identified as having high IgG reactivity to VAR2CSA by ELISA during assay optimisation. Seropositive samples were classified as having an OD greater than the mean + 3 standard deviations of malaria non-exposed Australian donors (n=15).

### Flow cytometry complement fixation assays with intact pRBCs

Pigmented trophozoite stage parasites at ∼10% parasitaemia and 0.5% haematocrit were incubated with 25% NS and 10% antibodies from PNG pregnant women or Australian malaria non-exposed donors for 1 hour at 37°C. pRBCs were then washed in 0.5% BSA in PBS and C1q, C3 and C5-9 detected by staining with polyclonal rabbit anti-human C1q, goat anti-human C3 or monoclonal mouse anti-human C5-9 antibodies followed by respective polyclonal Alexa-488 conjugated goat anti-rabbit, rabbit anti-goat or goat anti-mouse antibodies. pRBCs were detected by staining with ethidium bromide (Bio-Rad) at 1/1000 dilution. Levels of complement fixation on pRBCs were expressed as the Alexa-488 geometric mean fluorescent intensity of ethidium bromide stained pRBC populations. It is reported that aged RBCs can bind complement factors (46), and in assay work-up we found it was important to use fresh (recently collected) RBCs in assays. To prepare parasites for these assays, mature-pigmented trophozoites were purified by magnet filtration, mixed with fresh RBCs, and cultured for 48 hours to obtain a parasitaemia of ∼10% parasitaemia.

### Detection of complement fixation on pRBC surface by western blot

Magnet-purified pigmented trophozoite pRBCs (>95% purity) were incubated with a 10% pool of antibodies (N=9) from PNG pregnant women with high IgG reactivity to VAR2CSA-DBL5 (3D7) or a pool of antibodies (N=10) from malaria non-exposed Australian donors, and 25% NS as a source of complement, for 15 minutes at 37°C. pRBCs were then washed in cold PBS containing protease inhibitors. Samples were resuspended in reducing SDS sample buffer, heated at 95°C for 5 minutes and proteins separated on 4%-12% Bis-Tris gels (Invitrogen). Proteins were then transferred to nitrocellulose membranes (Invitrogen). C1q (29 Kd) was detected by staining with a rabbit anti-C1q antibody. C3b was detected using an anti-C3 antibody that labels both C3b and iC3b components that comigrate at ∼68kDa (21). Labelling of heat shock protein 70 (HSP-70) was used as a loading control.

### Assessment of complement-mediated killing of pRBCs

Mature pigmented trophozoite stage CS2 parasites at 0.5% parasitaemia and 3% haematocrit were cultured in complete medium in 96 well plates in the presence of 10% antibodies from PNG pregnant women (N=7) alone or with additional 25% NS or HIS. A negative control with no PNG or Australian malaria non-exposed donor antibodies and no NS or HIS added was also included in the assay. After approximately 24 hours incubation when parasites were at ring stages, samples were washed three times in RPMI to remove complement and antibodies and then returned to culture at 3% haematocrit in complete culture medium for a further 24 hours until parasites reached the mature pigmented trophozoite stage. Parasitaemia was measured by flow cytometry following staining with ethidium bromide at 1/1000 dilution. All samples were run blinded in triplicate and in 3 separate experiments.

### Statistical analyses

Statistical analyses were performed using STATA v13.1. Graphs were generated using GraphPad Prism v7. Continuous variables were compared using non-parametric Mann-Whitney U test or Wilcoxon matched-pairs signed rank tests. Correlations between continuous variables were assessed using Spearman’s rank correlation coefficient (rho). The association between antibody-mediated complement fixation and placental infection was tested using logistic regression with levels of complement fixing antibodies fitted as continuous variables. To aid interpretation, coefficients were presented representing the difference in outcome of a high responder (75^th^ percentile) or a medium responder (50^th^ percentile) compared to a low responder (25^th^ percentile). The model was adjusted for gravidity (primigravid/multigravid), location of residence (town/village), smoking (no/yes), middle upper arm circumference (as a marker of undernutrition in pregnancy), sex of newborn, haemoglobin levels at enrolment, South East Asian Ovalocytosis and α+thalassaemia (wild type vs heterozygous or homozygous). We tested for evidence of effect modification by *P. falciparum* peripheral infection at enrolment or gravidity on birth outcomes using log-likelihood ratio tests, with and without an interaction term. For all analysis P values less than 0.05 were considered statistically significant.

## RESULTS

### Antibodies from pregnant women promote complement fixation on placental binding pRBCs

We tested whether acquired antibodies from pregnant women from a malaria endemic PNG region could fix and activate complement on pRBCs. Trophozoite stage pRBCs of the placental binding *P. falciparum* isolate CS2 were opsonized with antibodies from 302 PNG women or 15 malaria non-exposed Australian donors and tested for their ability to fix complement factors C1q or C3 by flow cytometry. Antibodies from malaria-exposed pregnant PNG women mediated significantly higher levels of complement C1q (Figure 1A) and C3 (Figure 1B) fixation to the surface of CS2 pRBCs, compared to malaria non-exposed control donors (both P<0.001) (characteristics of the 302 women included in this study data are shown in Table 1). The prevalence of complement-fixing antibodies among pregnant women was 66% (200/302) for C1q and 51% (155/302) for C3 (Figure 1A-B). C1q fixation on pRBCs was moderately positively correlated with C3 fixation on pRBCs (Spearman r=0.36, P<0.001). C3 fixation was abolished when using heat-inactivated serum (HIS) (complement inactive) as a control, compared to normal serum (NS) (complement active) (P<0.001) (Figure 1C). C1q fixation on the surface of pRBCs was confirmed by Western blot in the presence of antibodies from PNG pregnant women compared to malaria non-exposed antibody pool (Figure 1D). Some C3 fixation on pRBCs was detected when using malaria non-exposed antibodies or unopsonized controls, but levels were much lower than those seen with the PNG antibodies pool (Figure 1D). A subset of samples was tested for complement fixation using an isolate from PNG (XIE), which is genetically distinct from CS2 (47). There was significantly higher C3 fixation among PNG pregnant women’s samples (N=35) compared to non-exposed controls (N=10) (P<0.001) (Supplementary Figure S1). Correlation between CS2 and XIE was 0.44, p=0.012.

**Table 1:** Study population characteristics by *P. falciparum* infection at enrolment

| Variable | Total<br>(N=302) | Uninfected at enrolment<br>(N=199) | Infected at enrolment<br>(N=103) |
| --- | --- | --- | --- |
| **Enrolment Demographics** |  |  |  |
| Age | 24 (21 – 28) | 24 (21 – 30) | 23 (20 – 27) |
| Gestational age (weeks) | 26 (22 – 28) | 26 (22 – 28) | 25 (22 – 29) |
| Gravidity |  |  |  |
| Primigravid | 115/302 (38%) | 66/199 (33%) | 49/103 (48%) |
| Multigravid | 187/302 (62%) | 133/199 (67%) | 54/103 (52%) |
| Middle upper arm circumference | 22 (21 – 23) | 23 (21 – 24) | 22 (22 – 23) |
| Smokes | 60/301 (20%) | 40/199 (20%) | 20/102 (20%) |
| **Infections at enrolment** |  |  |  |
| Peripheral *P. falciparum*<sup>a</sup> | 103/302 (34%) | 0/199 (0%) | 103/103 (100%) |
| **Infections at delivery** |  |  |  |
| Peripheral *P. falciparum*<sup>a</sup> | 32/288 (11%) | 22 /190 (12%) | 11/98 (11%) |
| **Placental histology<sup>b</sup>** |  |  |  |
| No infection | 81/233 (35%) | 64/154 (41) | 17/79 (21%) |
| Acute infection | 90/233 (39%) | 64/154 (39%) | 30/79 (38%) |
| Chronic infection | 42/233 (18%) | 21/154 (14%) | 21/79 (27%) |
| Past infection | 20/233 (8%) | 9/154 (6%) | 11/79 (14%) |
| **Haemoglobin at delivery** |  |  |  |
| Haemoglobin (g/dl) | 9.4 (8.2 – 10.3) | 9.4 (8.2 – 10.5) | 9.2 (8.2 – 10.2) |
| Severe anaemia (<8g/dl) | 68/285 (24%) | 45/188 (24%) | 23/97 (24%) |
| **Residence** |  |  |  |
| Town | 7/299 (2%) | 7/197 (4%) | 0/102 (0%) |
| Village | 292/299 (98%) | 190/197 (96%) | 102/102 (100%) |
| **Maternal Genetics** |  |  |  |
| **SAO** |  |  |  |
| Normal | 261/302 (86%) | 176/199 (88%) | 85/103 (83%) |
| SAO | 41/302 (14%) | 23/199 (12%) | 18/103 (17%) |
| **α<sup>+</sup>thalassaemia** |  |  |  |
| Normal α globin | 59/302 (20%) | 40/199 (20%) | 19/103 (19%) |
| Heterozygous | 112/302 (43%) | 72/199 (36%) | 40/103 (39%) |
| Homozygous | 131/302 (37%) | 87/199 (44%) | 44/103 (43%) |

Data are presented as median (25<sup>th</sup> percentile – 75<sup>th</sup> percentile) and n/total (%) for categorical variables693 <sup>a</sup>Parasite positive by light microscopy, confirmed by PCR. <sup>b</sup>Placental infection confirmed by histology.
Missing data for variables of; age and gestational age (n=13), smoker (n=1), middle upper arm
circumference (n=12), smoke (n=1), peripheral parasitaemia at delivery (n=14), placental histology
(n=69), Hb at delivery (n=17) and location of residence (n=3)

**Figure 1:**
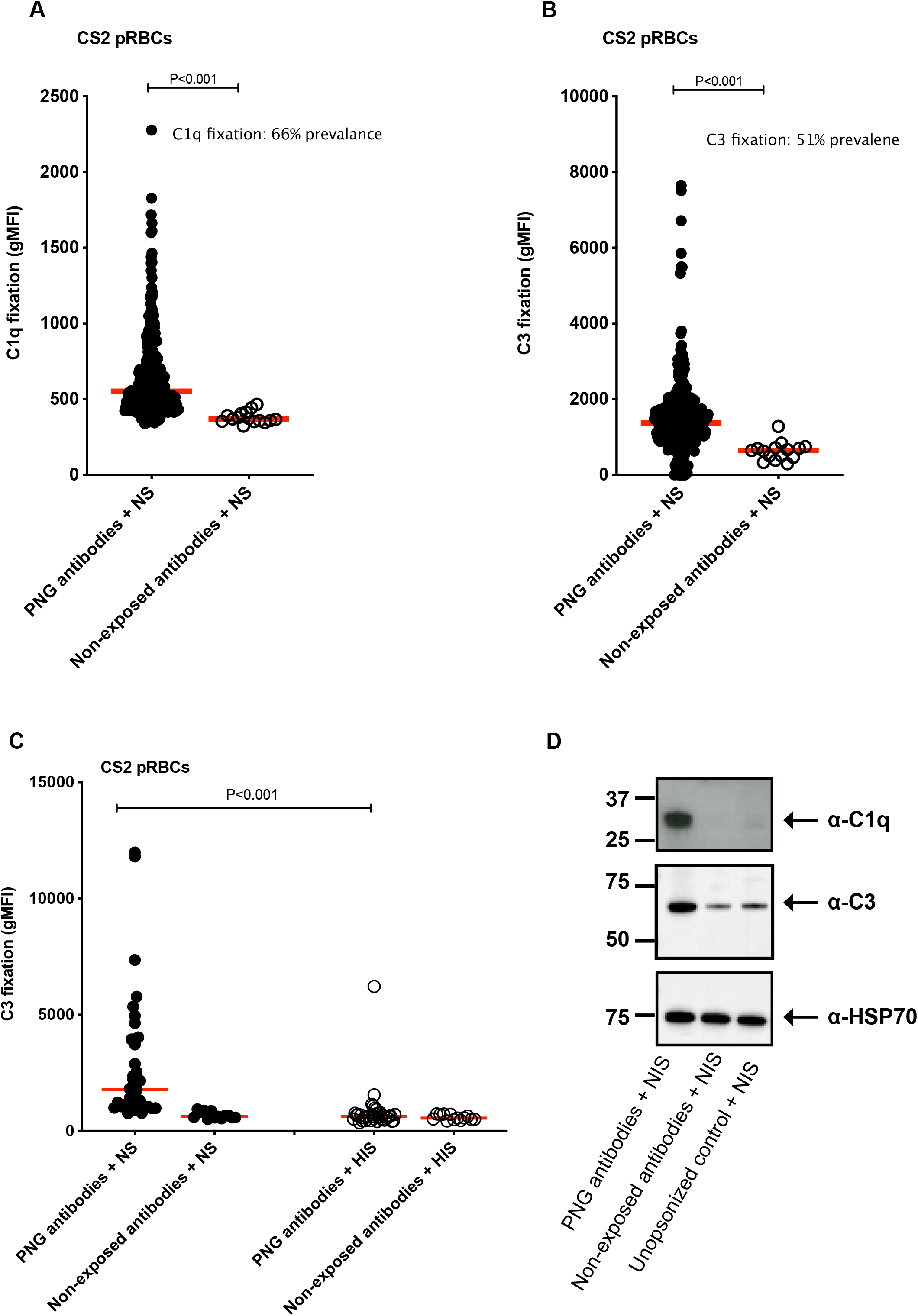
Antibodies from malaria-exposed PNG pregnant women mediated complement fixation on CS2 pRBCs. Complement **(A)** C1q and **(B)** C3 fixation on CS2 pRBCs by flow cytometry. Complement fixation was tested using antibodies from PNG pregnant women (n=302) and Australian malaria non-exposed donors (n=15) in the presence of complement (NS). **(C)** Complement C3 fixation on CS2 pRBCs in the presence of NS or HIS by flow cytometry. CS2 pRBCs were opsonised with antibodies from 35 randomly selected PNG pregnant women or Australian malaria non-exposed donors (N=13) and in the presence of either normal serum or heat inactivated serum. Complement fixation is presented as the average geometric mean fluorescence intensity (gMFI) of each sample tested in duplicate and the red line represents the median gMFI for all samples tested. **(D)** C1q and C3 fixation by Western blot using magnet purified CS2 pRBCs that were first opsonized with a pool of PNG or malaria non-exposed antibodies, in the presence of NS. Unopsonized sample (CS2 pRBCs incubated without antibody, but with NS) was used as a negative control and heat shock protein 70 (HSP70) as a loading control.

### Antibodies to placental binding parasites show limited MAC formation and activity

Using antibodies from a subset of women (selected on the basis of showing moderate to high C1q fixation (Supplementary Figure S2A-B)) we tested for evidence of C5-9 fixation and MAC activity on the surface of CS2 PRBCs. We observed no evidence of elevated C5-9 fixation on pRBCs by flow cytometry when these PNG antibodies were used to opsonize pRBCs in the presence of normal serum (Figure 2A). Additionally, we did not observe any significant correlation between C5-9 fixation and C1q or C3 fixation on the surface of CS2 pRBCs (Figure 2B). To assess whether there was significant MAC activity leading to pRBC lysis or impacting on pRBC viability, we tested whether incubation of CS2 pRBCs in the presence of PNG women’s antibodies (N=7) with active complement (NS) compared to heat-inactivated serum (inactive complement) impacted parasite replication *in vitro*. There was no evidence of reduced *in vitro* growth of pRBCs that were exposed to antibodies and active complement (Figure 2C). Therefore, these data suggest that while there is fixation of C1q and activation of C3 on the surface of pRBCs, there is limited formation of MAC or complement-mediated lysis of pRBCs.

**Figure 2:**
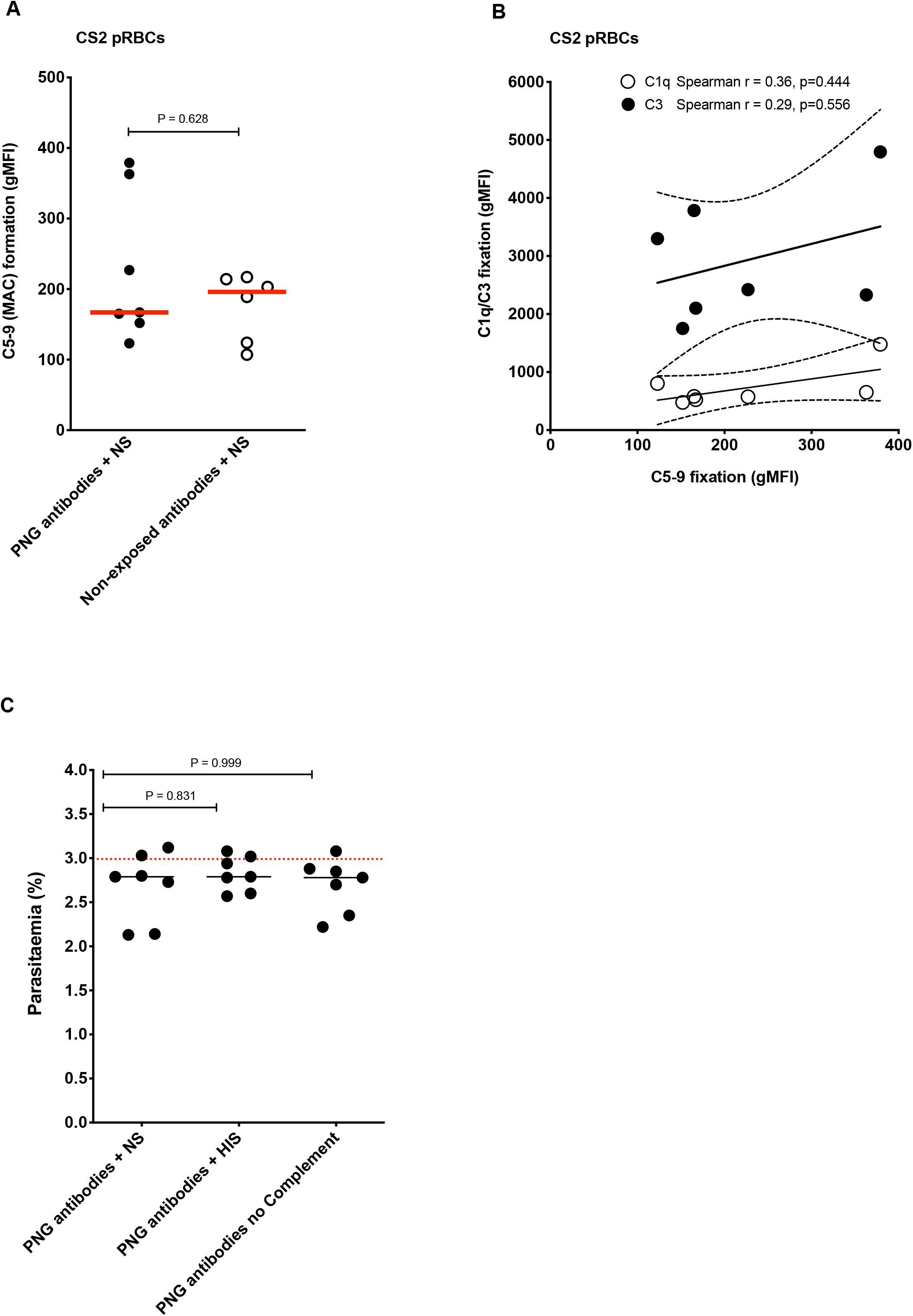
Limited evidence for complement C5-9 fixation and MAC activity on the surface of CS2 pRBCs. **(A)** Complement C5-9 fixation on CS2 pRBCs was tested by flow cytometry following opsonization with either antibodies from a subset of 7 PNG donor samples (with moderate to high C1q/C3 fixing activity on CS2 pRBCs) or 6 Australian malaria non-exposed donors in the presence of NS. Complement fixation is presented as the average gMFI of each sample tested in duplicate and the red line represents the median gMFI for all samples tested in each category. **(B)** Correlation between complement C5-9 fixation and C1q/C3 fixation tested on a subset of 7 PNG donor samples (with moderate to high C1q/C3 fixing activity on CS2 pRBCs). **(C)** Complement-mediated pRBC killing/lysis was examined by culturing CS2 pRBCs opsonized with antibodies from PNG pregnant women (n=7) in the presence of normal serum, heat-inactivated serum or no-added serum and measuring parasitaemia by flow cytometry after two growth cycles. Data are presented as the average parasitaemia of each sample run in triplicate over 3 independent experiments. The dotted red line represents the median parasitaemia of CS2 pRBCs cultured in the absence of either PNG or Australian malaria non-exposed antibodies and NS or HIS. Comparisons were made using Mann-Whitney U-test.

### PfEMP1 is a major target of complement fixing antibodies

PfEMP1 has been implicated as a major antigen on the surface of pRBCs (48). Therefore, we quantified its significance as a target of complement fixing antibodies. We first established that antibody samples from PNG pregnant women fixed C1q on recombinant VAR2CSA DBL5 (Figure 3A) and DBL3 domains (Figure 3B) at significantly higher levels compared to malaria non-exposed donors (both P<0.001); 57% (171/302) and 56% (170/302) of PNG samples were classified as positive for C1q fixation against DBL5 and DBL3 respectively. C1q fixation on DBL5 and DBL3 were moderately correlated with each other (Spearman r=0.63, P<0.001) and with C1q fixation on pRBCs (Spearman r=0.54 and r=0.50 respectively, P<0.001). C3 fixation on DBL5, tested for a subset of 35 randomly selected PNG antibody samples, was also significantly higher compared to malaria non-exposed donor samples (n=10) (P <0.001) (Figure 3C) and strongly correlated with C1q fixation (Spearman r=0.89, P<0.001) (Figure 3D). C3 fixation on DBL5 versus pRBCs were moderately correlated (Spearman r=0.48, P=0.003). Using a subset of 35 randomly selected PNG samples and 10 Australian malaria non-exposed donor samples, we confirmed that acquired antibodies fixed complement to a different allele of VAR2CSA, DBL5 domain (7G8) (Supplementary Figure S3). The correlation between complement C3 fixation to the 2 different alleles DBL5 (3D7) and DBL5 (7G8) was 0.61, p<0.001.

**Figure 3:**
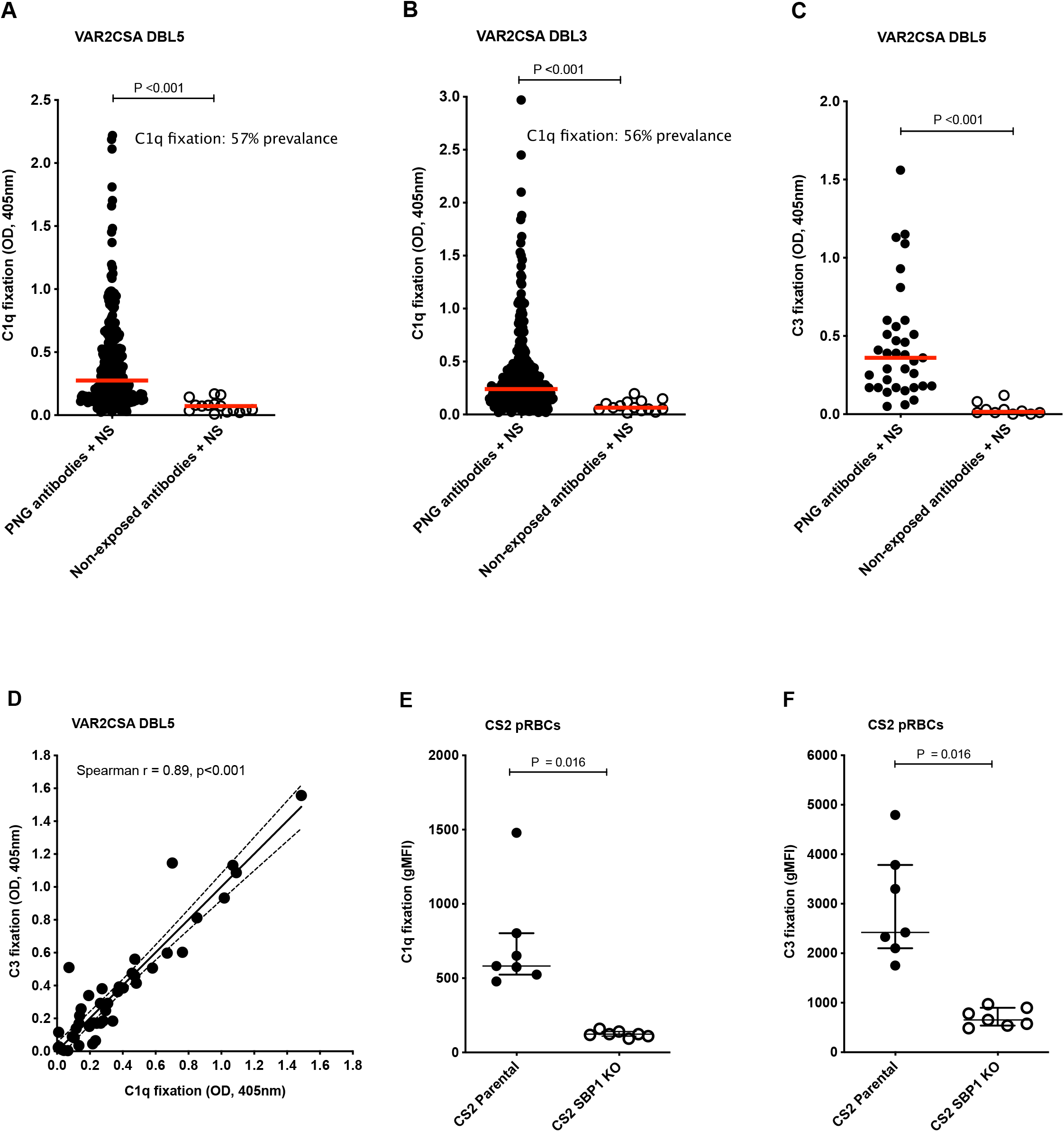
VAR2CSA PfEMP1 is a major target of complement fixing antibodies. Complement C1q fixation on **(A)** VAR2CSA-DBL5 (3D7) and **(B)** VAR2CSA-DBL3 (7G8) recombinant proteins using antibodies from 302 PNG pregnant women and 15 malaria non-exposed Australian donors. **(C)** C3 fixation on VAR2CSA-DBL5 (3D7) using antibodies from 35 randomly selected PNG pregnant women and 10 malaria non-exposed Australian donors (samples tested in duplicate). The median OD for all samples in each category is represented by the red line. Seropositivity was defined as OD > mean + 3 standard deviations of the 15 malaria non-exposed Australian controls. **(D)** Correlation between complement C1q and C3 fixation on VAR2CSA-DBL5 (3D7) using antibodies from 35 randomly selected PNG pregnant women (samples tested in duplicate). Complement **(E)** C1q and **(F)** C3 fixation on pRBCs was compared between the CS2-parental isolate and CS2 SBP1KO strain, which has reduced PfEMP1 surface expression. pRBCs were opsonized with 7 PNG antibody samples highly reactive to CS2 pRBCs. Complement fixation presented as the average gMFI of each sample run in duplicate from two independent experiments and bars show overall median gMFI and interquartile ranges.

We next evaluated antibody-mediated complement fixation on the pRBC surface using a CS2 *P. falciparum* isolate that was genetically-modified to reduce PfEMP1 surface expression; CS2 skeleton-binding protein 1 knockout (CS2-SBP1KO) (49). We found that C1q (Figure 3E) and C3 (Figure 3F) fixation were greatly reduced on the surface of CS2-SBP1KO pRBCs compared to CS2-parental pRBCs (P=0.016), indicating PfEMP1 as the major target of antibodies. Median C1q (Figure 3E) and C3 (Figure 3F) fixation was 76% and 53% lower, respectively, with CS2-SBP1KO pRBCs compared to CS2-parental pRBCs.

C1q fixation on VAR2CSA strongly correlated with IgG binding to both domains (DBL5; Spearman’s r=0.76, P<0.001, DBL3; Spearman’s r=0.75, P<0.001) in the PNG cohort (Table 2). VAR2CSA IgG subclass responses, tested on the random selection of 35 PNG samples, were predominantly IgG1 (80% seroprevalence) and IgG3 (37.1% seroprevalence) (Supplementary Figure S4) and moderately correlated with C1q fixation on DBL5 and DBL3 (Table 2).

**Table 2:** Spearman’s rank correlations between C1q fixation and antibodies to VAR2CSA DBL5 and DBL3 domains

| *Ig* | *C1q fixation* |  |
| --- | --- | --- |
|  | **DBL5** | **DBL3** |
| *Total IgG* | 0.76 | 0.75 |
| *IgG1* | 0.58 | 0.51 |
| *IgG2* | 0.46 | 0.35 |
| *IgG3* | 0.56 | 0.40 |

All correlations were statistically significant; P<0.001. IgG4 responses were not detectable in this cohort. N=302

### Antibody-mediated complement fixation is associated with gravidity and infection status and protection against placental infection

We found significantly higher C1q (Figure 4A; P<0.001) and C3 (Figure 4B; P=0.009) fixation on CS2 pRBCs among multigravid compared to primigravid women (Supplementary Figure S5A-B). Additionally, active peripheral *P. falciparum* infection at study enrolment was associated with higher complement C1q fixation on DBL5 (Figure 4G; P<0.001) and DBL3 domains (Figure 4H; P<0.001) compared to no infection at study enrolment (Supplementary Figure S5C-D).

**Figure 4:**
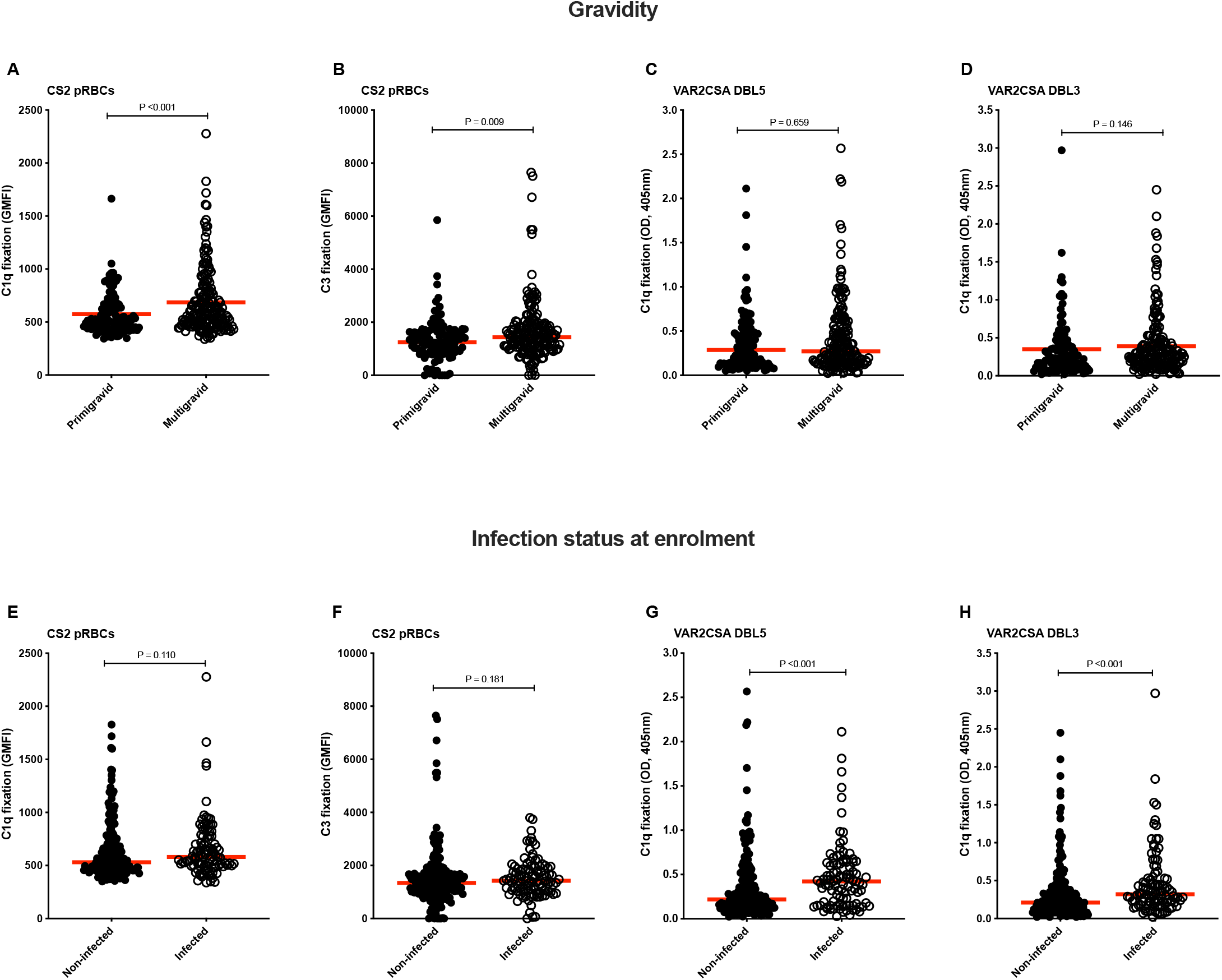
Complement fixation on VAR2CSA is associated with gravidity and *P. falciparum* peripheral infection status at enrolment. Complement C1q and C3 fixation was tested either on CS2 pRBCS by flow cytometry or on recombinant VAR2CSA-DBL5 (3D7) and VAR2CSA-DBL3 (7G8) domains by ELISA using samples from 302 PNG donors (primigravid n=115, multigravid n=187; non-infected=199, infected n=103). Complement **(A)** C1q and (**B)** C3 fixation on CS2 pRBCs by gravidity. Complement C1q fixation on **(C)** VAR2CSA-DBL5 (3D7) and (**D)** VAR2CSA-DBL3 (7G8) by gravidity. Complement **(E)** C1q and **(F)** C3 fixation on CS2 pRBCs by *P. falciparum* infection status at enrolment. Complement C1q fixation on **(G)** VAR2CSA-DBL5 (3D7) and **(H)** VAR2CSA-DBL3 (7G8) by *P. falciparum* infection status at enrolment. Data are presented as the average gMFI (flow cytometry) or optical density (OD; by ELISA) of each sample tested in duplicate and the red line represents the median gMFI or the OD. P values were calculated using the Mann-Whitney U test.

We tested for associations between antibody-mediated complement fixation and protection from placental malaria infection using logistic regression analysis. We tested the hypothesis that when a pregnant woman is infected with *P. falciparum*, those who generate higher levels of complement-fixing antibodies may be less likely to develop placental parasitemia, compared to those with low complement-fixing antibodies. Among women with *P. falciparum* infection at enrolment (median gestation 26 weeks; Table 1), those with high and intermediate levels of complement-fixing antibodies had significantly lower risk of placental infection when compared to those with low complement fixing antibodies (Figure 5A) (Table S1). Protective associations were observed with C1q or C3 fixation on placental-binding pRBCs, or recombinant VAR2CSA-DBL5 domain; associations were weaker for C1q-fixation on the DBL3 domain. Further analysis of placental malaria stratified by placental histology confirmed a reduced risk of acute and chronic placental infection with high and intermediate levels of complement-fixing antibodies (Tables S2 & S3). However, it is important to note that statistical power is reduced in these sub-group analyses. Among women who were not parasitemic at enrolment, there was no significant association between antibody complement fixation on pRBCs (C1q or C3) and risk of placental infection.

**Figure 5:**
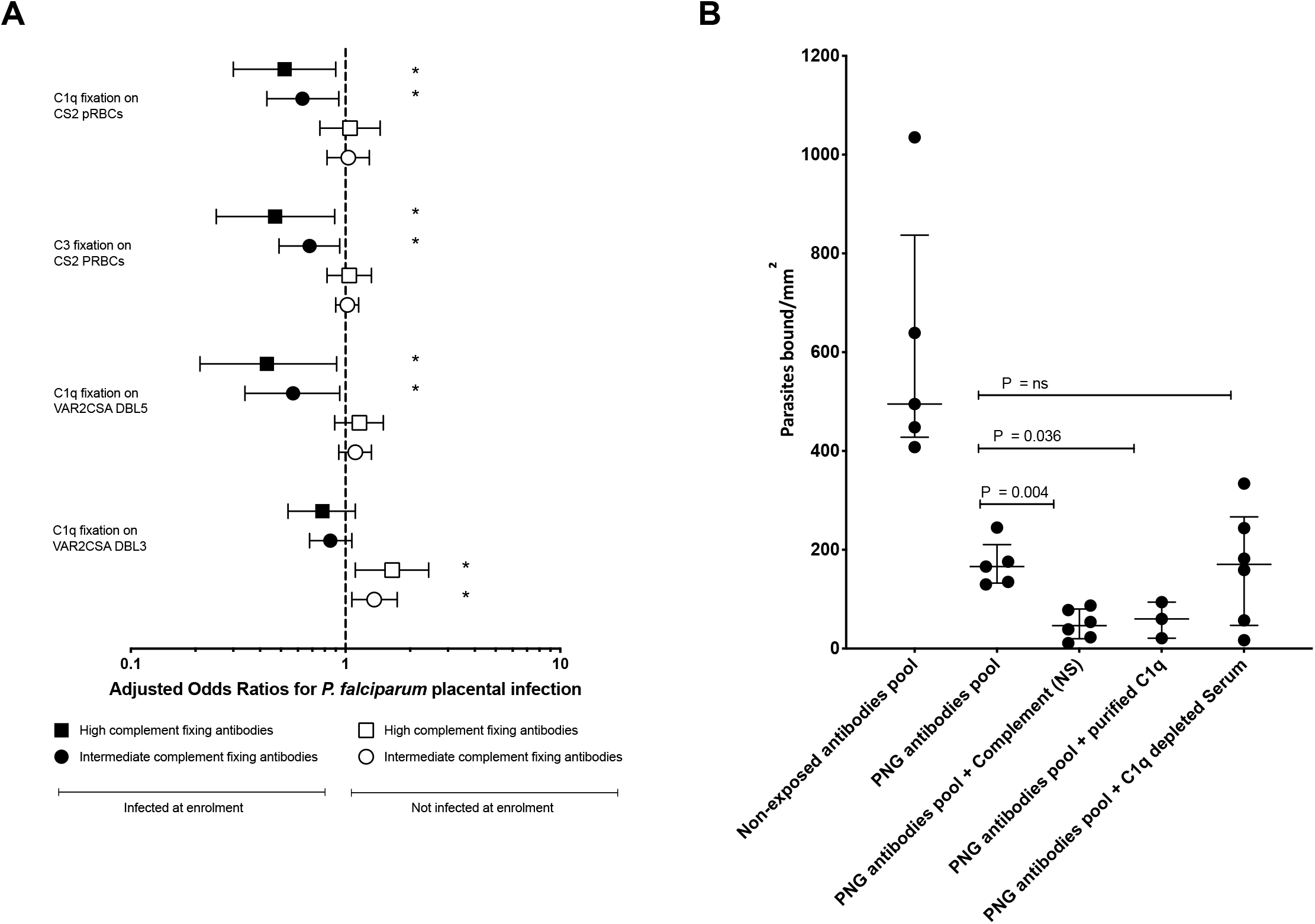
Complement fixing antibodies are associated with reduced risk of *P. falciparum* placental infection and enhanced inhibition of pRBC binding to CSA. **(A)** Odds ratios for *P. falciparum* placental infection for complement C1q and C3 fixation levels stratified by *P. falciparum* infection status at enrolment, using logistic regression. Model included complement fixation as a continuous dependent variable, effect modification by *P. falciparum* peripheral infection status at enrolment (uninfected vs infected). Model includes additional adjustment for gravidity (primigravid vs multigravid), location of residence (town vs village), smoking (no vs yes), middle upper arm circumference in cm, gender of newborn (male vs female), haemoglobin levels (g/dl) at enrolment, South East Asian Ovalocytosis (wild type vs SAO) and α+thalassaemia (wild type vs heterozygous/homozygous). A total of 140 uninfected and 71 *P. falciparum* infected (at enrolment ANC visit) individuals were included in the analysis of complement fixation on pRBCs and 141 uninfected and 71 *P. falciparum* infected individuals for complement fixation on VAR2CSA recombinant protein domains (VAR2CSA-DBL5 (3D7) & VAR2CSA-DBL3 (7G8)). Plots represent odds ratios for *P. falciparum* placental infection in individuals with high (75^th^ percentile) and intermediate (50^th^ percentile) complement fixing antibodies relative to low antibody levels (25^th^ percentile) and bars represent 95% confidence intervals. Statistically significant associations (P<0.05) are shown in asterisks. **(B)** CS2 pRBCs binding to CSA was tested following opsonisation of CS2 pRBCs with 10% PNG antibody pool in the presence of NS, purified human C1q or C1q-depleted human serum. A malaria non-exposed antibody pool was used as a negative control. Plots represent median and interquartile range of pRBCs bound to CSA per mm^2^. Each data point represents an independent assay with 3-6 independent assays performed for each condition.

### Antibody-mediated complement fixation enhances inhibition of pRBC adhesion to the placental receptor CSA

Complement-fixation on cell surfaces is known to enhance phagocytic clearance through interactions of C3b and C4b with numerous complement receptors expressed on cells such as monocytes and neutrophils (19). We investigated whether fixation of complement on pRBCs by antibodies might have an additional function of enhancing inhibition of pRBC binding to CSA. Using a pool of antibodies from pregnant PNG women, we found that antibodies, under all conditions, gave significant inhibition of binding of CS2 pRBCs to CSA compared to malaria non-exposed antibodies (Figure 5B) (P<0.05). In the presence of complement (NS), PNG antibodies significantly reduced binding of CS2 pRBCs to CSA (median binding 47 pRBCs/mm^2^, P=0.004) compared to PNG antibodies alone (166 pRBCs/mm^2^); reduction of 72% (Figure 5B). Since C1q is the first step in classical complement activation by antibodies, we evaluated C1q-depeted serum as a control, finding that PNG antibodies plus C1q-depleted serum had similar levels of binding to PNG antibodies alone (171 pRBCs/mm^2^, P>1.000), and significantly less inhibition than PNG antibodies with NS. Importantly, PNG antibodies, in the presence of human C1q only (not including other complement components) resulted in similar levels of binding inhibition as seen with whole serum (Figure 5B) (60 pRBCs/mm^2^, P=0.004). These findings suggest C1q is the major factor important in enhancing binding inhibition by antibodies (Figure 5B).

## DISCUSSION

Antibodies are thought to mediate protection against placental infection by *P. falciparum*. However, the mechanisms mediating immunity are not fully understood. Here, we reveal a new mechanism in immunity to malaria in pregnancy, which may contribute to reducing the risk of placental parasitemia. We show that acquired antibodies among pregnant women can mediate complement fixation on placental-binding *P. falciparum* pRBCs. Using genetically-modified *P. falciparum* pRBCs and recombinant antigens, we found that antibody-mediated complement-fixation predominantly targeted PfEMP1 (VAR2CSA) expressed on the surface of pRBCs. Importantly, higher complement-fixing antibodies were associated with a reduced risk of placental parasitaemia and resulted in enhanced inhibition of pRBC binding to CSA suggesting this mechanism contributes to immunity to MiP.

We demonstrated the ability of acquired antibodies to fix and activate complement using several approaches. Antibodies could fix C1q, the first step in the classical pathway and C3, indicating complement activation. This was demonstrated by labelling complement components on pRBCs by flow cytometry, and by western blotting of pRBC protein extracts, as well as using recombinant VAR2CSA domains. Complement fixation among antibodies from malaria-exposed pregnant women was significantly higher than antibodies from non-exposed donors and correlated with IgG reactivity. Furthermore, complement-fixing antibodies were generally higher among multigravid than primigravid women, consistent with the reported acquisition of immunity to MiP. Consistent with data from Africa (15, 16), IgG1 (80% seropositivity) and IgG3 (37% seropositivity) dominated responses in PNG women and moderately correlated with complement-fixing antibodies. We observed some complement C3 fixation when using malaria non-exposed donor antibodies or non-opsonized pRBCs, suggesting some activation via the antibody-independent alternate pathway; however, complement fixation was always much higher in the presence of antibodies from pregnant women supporting a greater role for the classical pathway. Significant polymorphisms occur in VAR2CSA, which can impact on binding of acquired IgG (47, 50, 51). Previously we established that the CS2 isolate used in this study is well recognised by malaria-exposed pregnant women in PNG, and IgG reactivity to CS2 pRBCs strongly correlated with IgG reactivity to a placental-binding clinical isolate from PNG (37).

Interestingly, there was no clear evidence of enhanced MAC formation on pRBCs by antibodies, even though antibodies promoted C1q and C3 fixation. Consistent with this, complement did not result in pRBC lysis or inhibition of growth. Complement regulatory proteins expressed on RBCs (e.g. CD59) likely prevent effective MAC formation, as suggested by older studies (26). Our findings highlight the complexities of adaptive humoral immunity. While host regulatory mechanisms designed to protect RBCs may prevent MAC formation and lysis of pRBCs, antibody-complement interactions can still mediate effects through C1q and C3 fixation in acquired immunity targeting pRBCs.

Two earlier studies showed evidence of complement fixation by antibodies among non-pregnant individuals on pRBCs with known binding to CD36 and ICAM-1 endothelial receptors (26, 27). However, the acquisition and targets of these antibodies were not assessed, nor were associations with protective immunity. In contrast to our findings, one recent study evaluated purified IgG from a pool of plasma samples from malaria-exposed pregnant Ghanaian women and a VAR2CSA monoclonal antibody (52). Complement fixation was detected on recombinant VAR2CSA, but not on the surface of pRBCs. However, the acquisition and functions of antibodies, or associations with clinical features or outcomes were not assessed. The detection of limited complement fixation on pRBCs in that study, compared to our results, may reflect differences in the sensitivity of different assays and the reagents used, and in that study the authors used 1% fresh serum as a source of complement. Additionally, using a pool of samples would contribute to a reduced ability to detect complement-fixing antibodies if the prevalence of such antibodies is low in the population.

Importantly, complement fixing antibodies at enrolment were prospectively associated with a reduced risk of placental parasitaemia at delivery, suggesting this immune mechanism may contribute to reducing parasitemia. Our further exploratory analysis of associations with active and chronic infection subgroups (defined by placental histology) found similar associations. This protective association was only evident among women with *P. falciparum* infection at baseline, suggesting that women who mount higher complement-fixing antibodies when challenged with infection more effectively control and clear infection. Among women without infection at enrolment, there was no significant association between complement fixing antibodies to pRBCs and risk of placental infection. Where malaria transmission is heterogenous only a proportion of the population are exposed to infection. One approach to address this is to stratify the analysis of associations with protection by infection status at baseline (53). Other studies investigating associations between antibodies and protection against malaria in children (54, 55) or pregnant women (56) have used this approach, revealing protective associations only in the infected group who have documented exposure to malaria. A potential limitation of our study is that women commenced in the study in mid-pregnancy (most were second trimester), even though we recruited pregnant women at their first antenatal clinic visit. Presenting to first antenatal clinic in the second trimester is commonly observed in PNG (57, 58). This means that we cannot fully account for malaria exposure history in their pregnancy. A further limitation is that placental tissue was available only for 77% women. However, we did not observe major difference in demographic or clinical features of women who did or did not have placental tissue collected (29). Ideally, a future study would enrol women early in pregnancy with frequent follow-up during pregnancy and employ a larger sample size to further investigate associations we observed here, including the relative roles of different antibodies and the influence of gravidity.

We found that complement significantly enhanced inhibition of pRBC adhesion to CSA by acquired antibodies, suggesting a potential mechanism mediating protective effects of complement-fixing antibodies. Additionally, it is well established that complement enhances phagocytosis through interactions with complement receptors expressed on monocytes and neutrophils, particularly via C3b (19, 59). Therefore, higher complement fixation by antibodies is likely to contribute to enhanced clearance of pRBCs. Anti-VAR2CSA antibodies have been proposed to protect against MiP possibly through inhibition of *P. falciparum* binding in the placenta along with enhanced phagocytosis (11, 15, 60, 61). Previous studies, however, have largely examined these functional mechanisms in the absence of complement. The initial step of the classical pathway of complement activation involving C1q appeared essential and sufficient in mediating inhibition enhancement. However, it should be noted that there is evidence that C1q can bind directly to CSA (62), emphasising the need for appropriate controls when assessing this effect. Others have shown complement can enhance inhibition of *P. falciparum* merozoite and sporozoite invasion (21, 23) and phagocytosis by monocytes (59). Prior studies of viruses have shown that binding of C1q by antibodies can enhance virus neutralization, without the need for additional complement components (20, 63). In contrast to protective roles of antibody-complement interactions, widespread complement activation has also been implicated in MiP pathology. Higher plasma C5a was associated with adverse birth outcomes including low birth weight, fetal growth restriction and preterm birth (28). Excess C5a is thought to mediate pathogenesis via inflammation and skews angiogenic profiles critical for normal placental vascularization and development towards an anti-angiogenic profile that promotes fetal growth restriction (28). Therefore, there is a balance between antibody-mediated complement activity on the surface of pRBCs that may be protective, versus widespread systemic complement activation that may be detrimental (64). We propose that acquisition of antibodies with the right specificity and properties leads to effective complement-fixing activity on the pRBC surface that reduces placental infection through mechanisms such as pRBC placental binding inhibition and enhanced phagocytic clearance. However, in the absence of protective antibodies, infection goes unchecked leading to high-density placental infections triggering excessive complement activation contributing to poor birth outcomes. Further studies to understand protective versus detrimental responses would be valuable.

While *P. falciparum* expresses multiple antigens on the surface of pRBCs that can be targeted by antibodies (48), we found that complement-fixing antibodies predominantly target PfEMP1. We used genetically-modified CS2 *P. falciparum* in which PfSBP1 has been disrupted to inhibit PfEMP1 surface expression (36). That there was still some complement fixation on pRBCs lacking PfEMP1 expression suggests that other surface expressed antigens might be secondary targets of complement-fixing antibodies, and these may include RIFINs and STEVORs (65), and warrant future investigation. C3 fixation against CS2 SBP1KO pRBCs might also be the result of complement activation via the antibody-independent alternate pathway. We further demonstrated that VAR2CSA is a target of complement-fixing antibodies using recombinant DBL3 and DBL5 domains, which are prominent targets of acquired antibodies associated with protection from MiP (40, 66). It would be valuable to investigate the complement fixing activity of antibodies induced by vaccines based on VAR2CSA in recent clinical trials (14, 67).

In conclusion, we have generated significant new evidence supporting antibody-complement interactions in immunity against MiP. These findings provide novel insights into the mechanisms mediating immunity in pregnancy and inform approaches to develop vaccines or other interventions against MiP.

## Data Availability

The datasets used and/or analysed during the current study are available from the
corresponding author on reasonable request.

## DECLARATIONS

### Ethics and approval of consent to participate

This study received ethical approval from the PNG Medical Research Advisory Council, the Melbourne Health Human Research Ethics Committee and Alfred Health Human Research Ethics Committee. Eligible women were read a statement describing the study and gave a written informed voluntary consent.

### Availability of data and materials

The datasets used and/or analysed during the current study are available from the corresponding author on reasonable request.

### Consent for publication

Not applicable. No details, images, or videos relating to an individual person are included.

### Author’s contributions

JGB, FJIF, SJR, and IM designed the research. DHO, MJB, AM, LR, JAC, DIS and AU performed the research, DHO and AM analysed the data and DHO and JGB wrote the manuscript with input from all authors. All authors read and approved the final version of the manuscript.

## Acknowledgements

The authors thank the mothers and their families for participation in this study, the staff of Alexishafen Health Centre for their enthusiastic cooperation, and staff of the PNG Institute for Medical Research, particularly Francesca Baiwog (now deceased), Prof. Peter Siba and Prof. Willie Pomat. We also thank Prof. Joseph Smith (Seattle Children’s Research Institute, Seattle, USA) for his generous contribution of all the VAR2CSA recombinant proteins and Dr. Paul Gilson for the anti-HSP70 antibody.

