## Supplementary tables and figures for "Reduced risk of placental parasitaemia associated with complement fixation on *Plasmodium falciparum* by antibodies among pregnant women"

**SUPPLEMENTARY MATERIAL**

### TABLES

**Table S1: Associations between complement fixation and placental malaria by infection status at enrolment**

| Complement fixation | | aOR (95% CI); p value | | |
|---|---|---|---|---|
| | | **Infected at enrolment** | **Uninfected at enrolment** | **lrt chi2 (p value)** |
| **CS2 pRBCs – C1q** | Low vs Medium | **0.82 (0.69-0.97); 0.021** | 1.01 (0.92-1.12); 0.792 | |
| | Low vs High | **0.52 (0.30-0.90); 0.021** | 1.05 (0.76-1.45); 0.792 | 5.54 (0.019) |
| **CS2 pRBCs – C3** | Low vs Medium | **0.70 (0.52-0.95); 0.021** | 1.02 (0.91-1.15); 0.774 | |
| | Low vs High | **0.47 (0.25-0.89); 0.021** | 1.04 (0.82-1.32); 0.774 | 5.76 (0.016) |
| **VAR2CSA-DBL5 – C1q** | Low vs Medium | **0.77 (0.60-0.97); 0.028** | 1.05 (0.97-1.14); 0.266 | |
| | Low vs High | **0.43 (0.21-0.91); 0.028** | 1.16 (0.89-1.50); 0.266 | 7.74 (0.005) |
| **VAR2CSA-DBL3 – C1q** | Low vs Medium | 0.91 (0.79-1.04); 0.167 | **1.21 (1.04-1.40); 0.013** | |
| | Low vs High | 0.78 (0.54-1.11); 0.167 | **1.65 (1.11-2.44); 0.013** | 9.35 (0.002) |

aOR - Adjusted odds ratios (aOR) for placental infection (acute and chronic). lrt – Likelihood ratio test comparing model with interaction term between *P. falciparum* peripheral infection status at enrolment and complement fixation to model without interaction term. Statistically significant associations (P<0.05) are shown in bold.

**Table S2: Associations between complement fixation and acute placental infection by infection status at enrolment**

| **Complement fixation** | | **aOR (95% CI); p value** | | |
|---|---|---|---|---|
| | | **Infected at enrolment** | **Uninfected at enrolment** | **lrt chi2 (p value)** |
| **CS2 pRBCs – C1q** | Low vs Medium | **0.78 (0.63-0.98); 0.030** | 1.03 (0.92-1.14); 0.638 | |
| | Low vs High | **0.45 (0.22-0.93); 0.030** | 1.09 (0.77-1.54); 0.638 | 6.54 (0.011) |
| **CS2 pRBCs – C3** | Low vs Medium | **0.65 (0.43-0.98); 0.037** | 0.99 (0.87-1.13); 0.876 | |
| | Low vs High | **0.41 (0.17-0.95); 0.037** | 0.98 (0.74-1.29); 0.876 | 4.15 (0.042) |
| **VAR2CSA-DBL5 – C1q** | Low vs Medium | 0.80 (0.60-1.07); 0.129 | 1.11 (1.00-1.24); 0.061 | |
| | Low vs High | 0.49 (0.19-1.23); 0.129 | 1.38 (0.99-1.93); 0.061 | 4.76 (0.029) |
| **VAR2CSA-DBL3 – C1q** | Low vs Medium | 0.82 (0.62-1.07); 0.146 | **1.18 (1.03-1.36); 0.021** | |
| | Low vs High | 0.58 (0.28-1.21); 0.146 | **1.55 (1.07-2.26); 0.021** | 5.80 (0.016) |

aOR - Adjusted odds ratios (aOR) for acute placental infection. lrt – Likelihood ratio test comparing model with interaction term between *P. falciparum* peripheral infection status at enrolment and complement fixation to model without interaction term. Statistically significant associations (P<0.05) are shown in bold.

**Table S3: Associations between complement fixation and chronic placental infection by infection status at enrolment**

| Complement fixation | aOR (95% CI); p value | | | |
|---|---|---|---|---|
| | | Infected at enrolment | Uninfected at enrolment | lrt chi2 (p value) |
| **CS2 pRBCs – C1q** | Low vs Medium | **0.64 (0.44-0.94); 0.023** | 1.00 (0.82-1.22); 0.978 | |
| | Low vs High | **0.24 (0.07-0.82); 0.023** | 0.99 (0.52-1.88); 0.978 | 4.86 (0.027) |
| **CS2 pRBCs – C3** | Low vs Medium | 0.51 (0.24-1.06); 0.071 | 1.10 (0.92-1.31); 0.281 | |
| | Low vs High | 0.24 (0.05-1.13); 0.071 | 1.23 (0.85-1.77); 0.281 | 5.11 (0.024) |
| **VAR2CSA-DBL5 – C1q** | Low vs Medium | 0.65 (0.40-1.06); 0.084 | 1.04 (0.86-1.25); 0.674 | |
| | Low vs High | 0.26 (0.06-1.20); 0.084 | 1.13 (0.64-2.02); 0.674 | 3.23 (0.073) |
| **VAR2CSA-DBL3 – C1q** | Low vs Medium | 1.07 (0.85-1.35); 0.557 | 1.22 (0.93-1.60); 0.143 | |
| | Low vs High | 1.20 (0.66-2.19); 0.557 | 1.69 (0.84-3.43); 0.143 | 0.52 (0.469) |

aOR - Adjusted odds ratios (aOR) for chronic placental infection. lrt – Likelihood ratio test comparing model with interaction term between *P. falciparum* peripheral infection status at enrolment and complement fixation to model without interaction term. Statistically significant associations ($P<0.05$) are shown in bold.

**Table S4: Associations between complement fixation and past placental infection by infection status at enrolment**

| **Complement fixation** | | **aOR (95% CI); p value** | | |
|---|---|---|---|---|
| | | **Infected at enrolment** | **Uninfected at enrolment** | **lrt chi2 (p value)** |
| **CS2 pRBCs – C1q** | Low vs Medium | 1.31 (0.85-2.02); 0.220 | 1.09 (0.94-1.26); 0.269 | |
| | Low vs High | 2.32 (0.60-8.92); 0.220 | 1.29 (0.82-2.04); 0.269 | 3.37 (0.066) |
| **CS2 pRBCs – C3** | Low vs Medium | 1.04 (0.87-1.25); 0.661 | 1.00 (0.66-1.51); 0.989 | |
| | Low vs High | 1.11 (0.69-1.81); 0.661 | 0.99 (0.33-2.94); 0.989 | 0.06 (0.805) |
| **VAR2CSA-DBL5 – C1q** | Low vs Medium | 0.75 (0.47-1.19); 0.215 | 1.10 (0.91-1.32); 0.339 | |
| | Low vs High | 0.39 (0.09-1.73); 0.215 | 1.35 (0.73-2.47); 0.339 | 0.74 (0.391) |
| **VAR2CSA-DBL3 – C1q** | Low vs Medium | 1.04 (0.70-1.54); 0.839 | 1.11 (0.84-1.46); 0.481 | |
| | Low vs High | 1.08 (0.48-2.46); 0.839 | 1.23 (0.69-2.20); 0.481 | 0.04 (0.846) |

aOR - Adjusted odds ratios (aOR) for past placental infection. lrt – Likelihood ratio test comparing model with interaction term between *P. falciparum* peripheral infection status at enrolment and complement fixation to model without interaction term. Statistically significant associations (P<0.05) are shown in bold.

**FIGURES**

**Figure S1: Antibodies from pregnant women promote complement fixation on the XIE *P. falciparum* isolate**

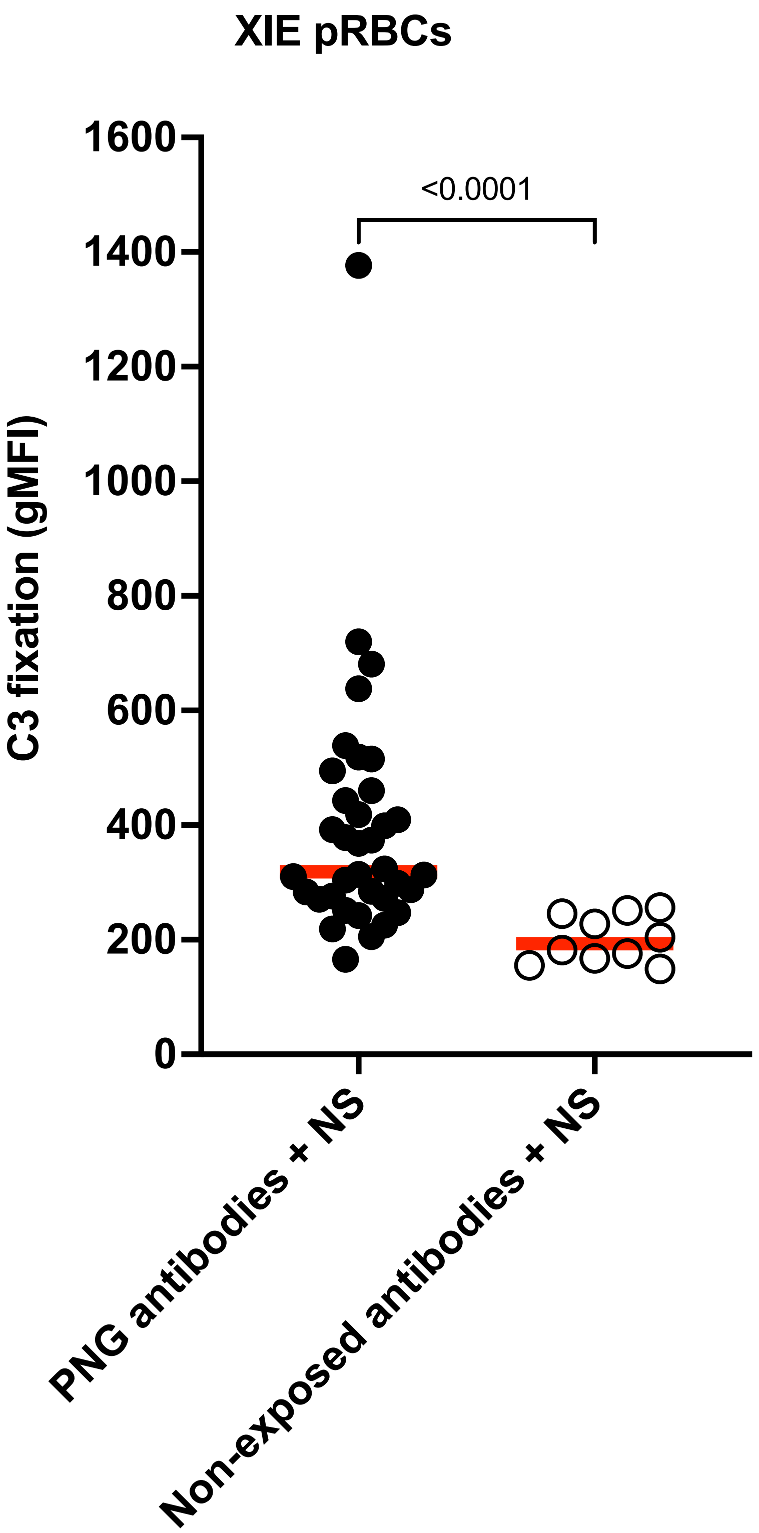

**Figure S2: C1q and C3 fixation on the surface of CS2 pRBCs for samples used to test for C5-9 fixation and MAC activity**

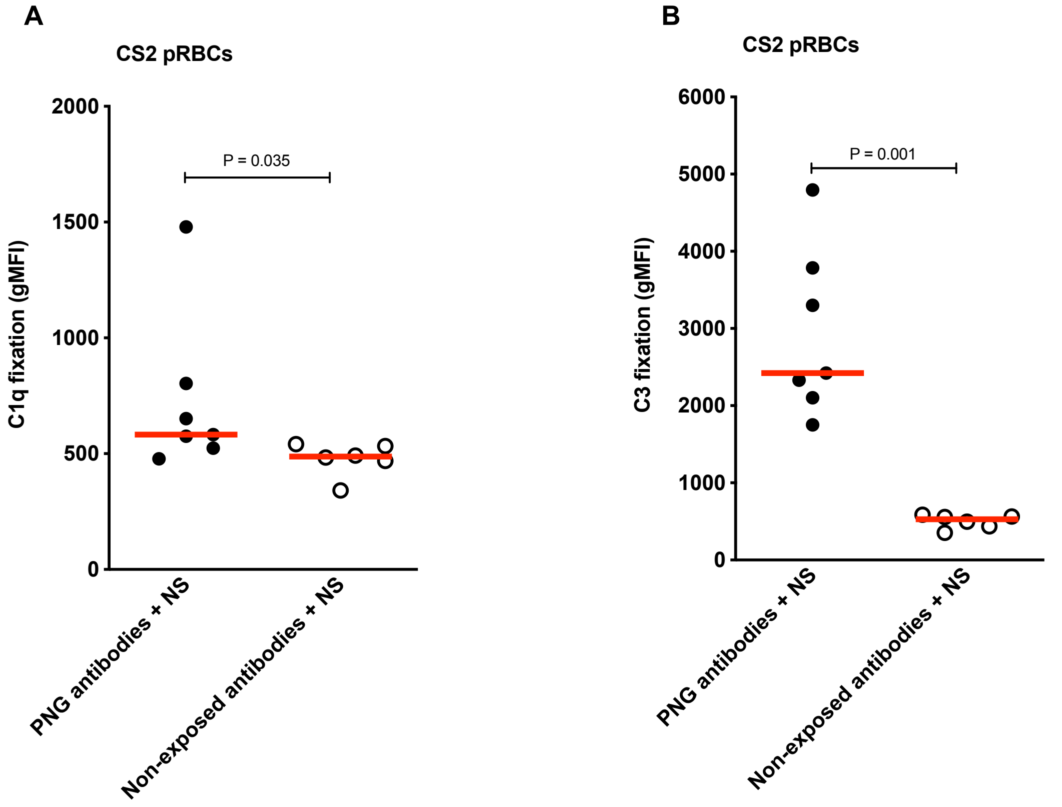

**Figure S3: Antibodies from pregnant women promote complement fixation on VAR2CSA DBL5 (7G8) recombinant protein**

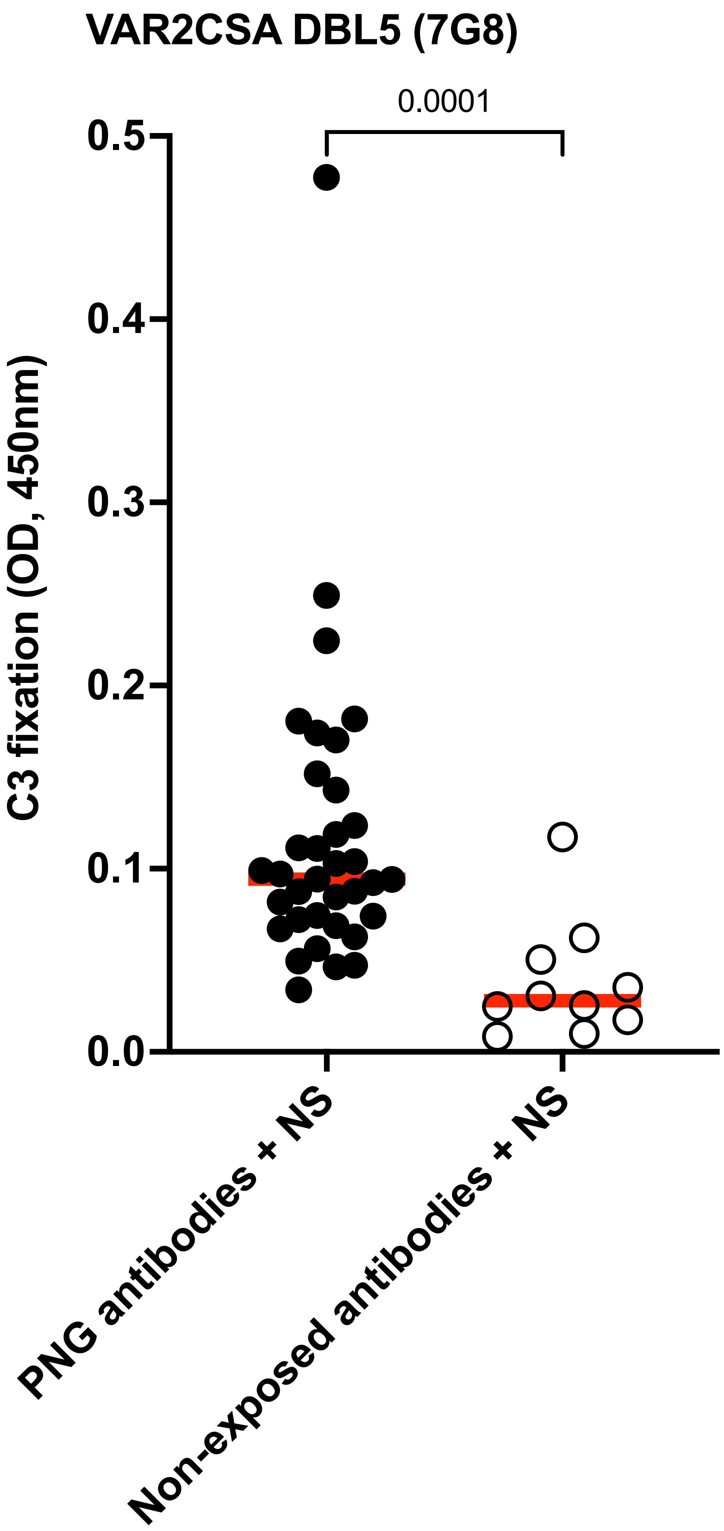

**Figure S4: Anti-VAR2CSA IgG subclass responses are predominantly IgG1 and IgG3**

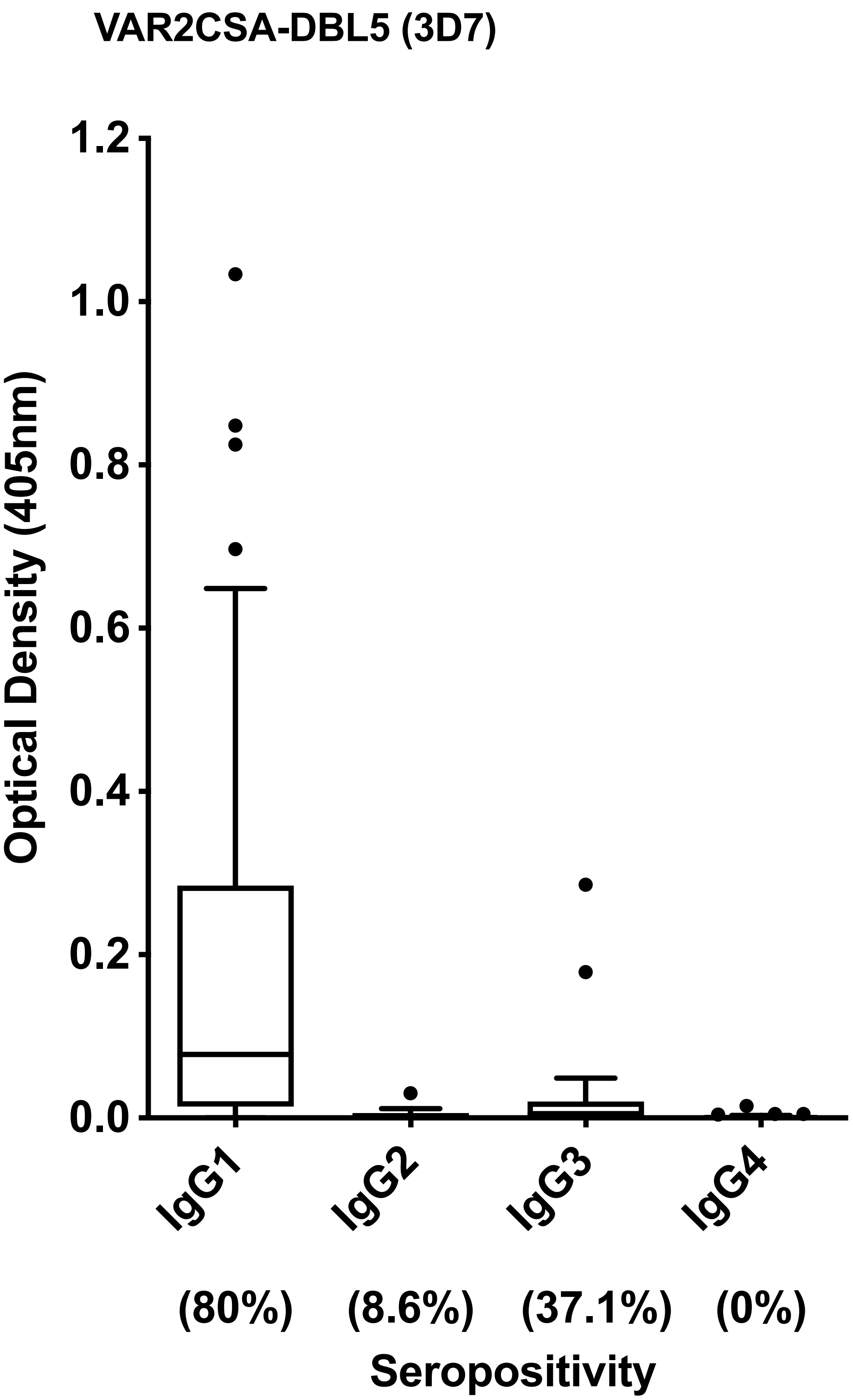

**Figure S5: Antibody-mediated complement fixation is associated with gravidity and infection status at enrolment**

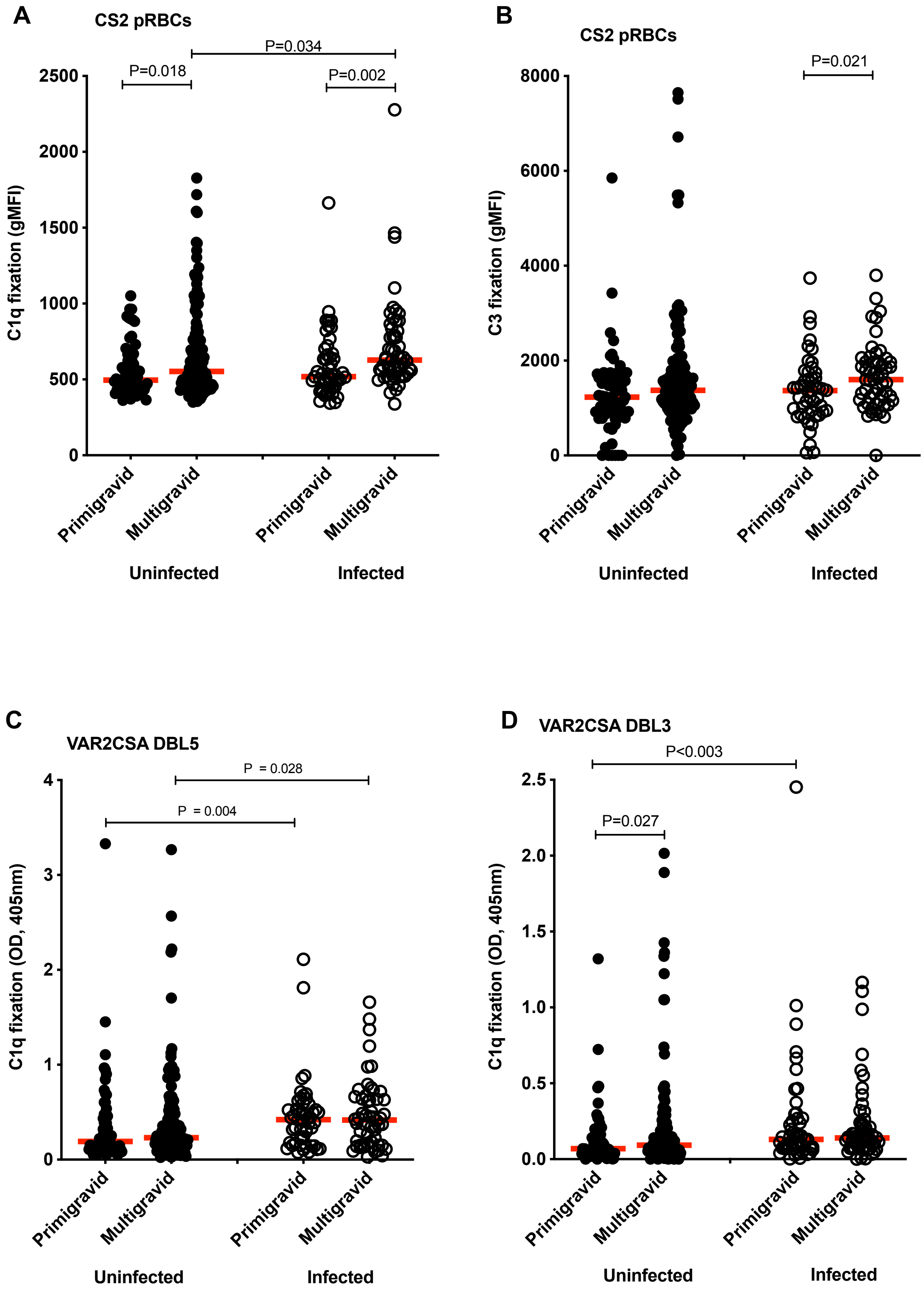

### FIGURE LEGENDS

**Figure S1: Antibodies from pregnant women promote complement fixation on the XIE *P. falciparum* isolate**

Complement C3 fixation on XIE pRBCs was tested in the presence of NS by flow cytometry on a subset of antibodies from 35 PNG pregnant women and 10 Australian malaria non-exposed donors. Complement fixation is presented as the average gMFI of each sample tested in duplicate and the red line represents the median gMFI for all samples tested in each category.

**Figure S2: C1q and C3 fixation on the surface of CS2 pRBCs for samples used to test for C5-9 fixation and MAC activity**

Complement **(A)** C1q and **(B)** C3 fixation on CS2 pRBCs was tested in the presence of NS by flow cytometry on a subset of antibodies from 7 PNG pregnant women (with moderate to high C1q/C3 fixing activity on CS2 pRBCs) or 6 Australian malaria non-exposed donors that were also tested for C5-9 fixation (Figure 2A). Complement fixation is presented as the average gMFI of each sample tested in duplicate and the red line represents the median gMFI for all samples tested in each category.

**Figure S3: Antibodies from pregnant women promote complement fixation on VAR2CSA-DBL5 (7G8) recombinant protein**

C3 fixation on VAR2CSA-DBL5 (7G8) was tested by ELISA using antibodies from 35 randomly selected PNG pregnant women and 10 malaria non-exposed Australian donors (samples tested in duplicate). The median OD for all samples in each category is represented by the red line.

**Figure S4: Anti-VAR2CSA IgG subclass responses are predominantly IgG1 and IgG3**

We tested for IgG subclass responses (IgG1, IgG2, IgG3 and IgG4) to VAR2CSA-DBL5 (3D7) by ELISA using a random selection of antibodies from 35 PNG pregnant women. Following incubation of antibodies to VAR2CSA-DBL5 (3D7) coated plates, samples were probed with anti-human IgG1, IgG2, IgG3 & IgG4 antibodies and results presented as optical density. Median values are represented by the horizontal line and boxes represent interquartile ranges, while dotted points represent outliers. Seropositivity is defined as having an OD greater than the mean + 3 standard deviations of malaria non-exposed Australian donors.

**Figure S5: Antibody-mediated complement fixation is associated with gravidity and infection status at enrolment**

**(A)** Complement C1q fixation on CS2 pRBCs by gravidity and *P. falciparum* infection status at enrolment. **(B)** Complement C3 fixation on pRBCs by gravidity and *P. falciparum* infection status at enrolment. **(C)** Complement C1q fixation on VAR2CSA-DBL5 (3D7) domain by gravidity and *P. falciparum* infection status at enrolment. **(D)** Complement C1q fixation on VAR2CSA-DBL3 (7G8) domain by gravidity and *P. falciparum* infection status at enrolment. Complement C1q and C3 fixation was tested either on CS2 pRBCS by flow cytometry or on recombinant VAR2CSA-DBL5 and VAR2CSA-DBL3 domains by ELISA using antibodies from 302 PNG donors (primigravid uninfected n=66, multigravid uninfected n=133, primigravid infected n=49, multigravid infected n=54)**.** Data are stratified by gravidity and *P. falciparum* infection status at enrolment and presented either as the as the average gMFI (flow cytometry) or OD (ELISA) of each sample run in duplicate with the median gMFI or OD for all samples in each category represented by the red line. Statistically significant P values (<0.05) are indicated in the plots.
